# Accents Still Confuse AI: Systematic Errors in Speech Transcription and LLM-Based Remedies

**DOI:** 10.1101/2025.08.29.25333548

**Authors:** Yasaman Fatapour, Jamil S. Samaan, Inclusive AI Research Group, Nicholas P Tatonetti

**Affiliations:** Department of Computational Biomedicine, Cedars-Sinai Medical Center, Los Angeles, California, USA; Karsh Division of Gastroenterology and Hepatology, Cedars-Sinai Medical Center, Los Angeles, California, USA; Department of Biomedical Engineering, University of California, Irvine, Irvine, California, USA; Cedars-Sinai Cancer, Cedars-Sinai Medical Center, Los Angeles, California, USA; Keck School of Medicine, University of Southern California, Los Angeles, California, USA; Department of Internal Medicine, Martin Luther King Jr. Community Hospital, Los Angeles, California, USA; Division of Cardiology, Department of Medicine, University of California, Irvine Medical Center, Orange, California, USA; Department of Medicine, Cedars-Sinai Medical Center, Los Angeles, CA, USA; Division of Gastroenterology and Hepatology, Department of Medicine, Bangkok Hospital Pattaya, Chonburi, Thailand; Department of Biomedical Informatics, Columbia University, New York, NY, United States; Department of Biomedical Informatics and Medical Education, University of Washington, Seattle, WA, United States

## Abstract

Accurate and timely documentation in the electronic health record (EHR) is essential for delivering safe and effective patient care. AI-enabled medical tools powered by automatic speech recognition (ASR) offer to streamline this process by transcribing clinical conversations directly into structured notes. However, a critical challenge in deploying these technologies at scale is their variable performance across speakers with diverse accents, which leads to transcription inaccuracies, misinterpretation, and downstream clinical risks. We measured transcription accuracy of Whisper and WhisperX on clinical texts across native and non-native English speakers and found that both models have significantly higher errors for non-native speakers. Fortunately, we found that post-processing the transcripts using GPT-4o recovers the lost accuracy. Our findings indicate that using a chained model approach, WhisperX-GPT, will enhance transcription quality significantly and reduce errors associated with accented speech. We make all code, models, and pipelines freely available.

## Introduction

There has been a marked acceleration in the development and application of artificial intelligence (AI) systems within healthcare in recent years. AI-driven technologies have been developed and introduced across the clinical continuum including decision support, clinical documentation, and patient engagement tools. (1–3) These systems are designed to support both healthcare professionals and patients, with the aim of improving care delivery, streamlining workflows, and enhancing health outcomes. Automatic speech recognition (ASR) technology forms the foundation of audio transcription within AI systems, enabling the conversion of spoken language into structured, machine-readable text. ASR models often serve as a critical bridge between AI systems and their human users by enabling seamless communication in clinical workflows. Despite their growing utilization, concerns have been raised regarding the equitable performance of ASR models across diverse user populations. (4–6)

ASR models process audio by segmenting speech into discrete units, extracting acoustic features, and applying statistical models to predict corresponding text. These models are further refined by language models that assign probabilities to likely word sequences, enabling contextual interpretation. While these approaches have achieved remarkable progress in many domains, their performance may vary when confronted with linguistic variation, including differences in accent, intonation, and grammatical structure. Studies have found ASR models often exhibit reduced transcription accuracy when processing speech from non-native English speakers, attributable to variations in pronunciation, syntax, and prosody. (7,8) We observed these errors first hand when evaluating an AI-scribe for endoscopic procedural notes and hypothesized that this problem may be a challenge in across healthcare settings. Such errors would have important implications for healthcare, where precise communication between AI systems and users is critical for safe and effective implementation.

Characterizing and addressing this disparity in ASR performance is critical to ensuring equitable implementation of AI technologies within the healthcare system. The healthcare workforce in the United States is notably diverse, with an estimated 18% of healthcare workers being foreign-born. (9) Furthermore, data from the U.S. Census Bureau indicate that the number of non-English speakers in the United States has been steadily increasing over the past four decades. (10)

In this pilot study, we evaluated the performance of two ASR models, Whisper and WhisperX, in transcribing standardized clinical audio from both native and non-native English speakers, including physicians and non-clinical participants. We found that both models performed significantly worse for non-native speakers, with Whisper more greatly affected. To reduce these disparities, we developed a post-processing pipeline using a large language model (LLM) and found significant reduction in these errors. Our results demonstrate that combining ASR with LLM correction enhances the reliability of AI-generated clinical documentation, particularly for diverse speaker populations. To our knowledge, this is the first study to systematically examine transcription bias based on language background and medical expertise, an essential step toward equitable AI integration in clinical workflows.

## Materials and Methods

We conducted a cross-sectional study of 20 participants who were stratified into four groups based on two binary variables: whether they were native English speakers and whether they had formal medical training. Native English speakers were defined as individuals who had completed formal education beginning in the sixth grade or earlier in the United States. Formal medical training was defined as current or prior enrollment in an accredited medical education program, including medical students, residents, fellows, or independently practicing physicians. This stratification resulted in four distinct groups each with 5 participants:

(1) Native English speakers with formal medical training
(2) Native English speakers without medical training
(3) Non-native English speakers with formal medical training
(4) Non-native English speakers without medical training.

We developed four standardized medical texts that represented typical clinical scenarios for the participants to read:

- TEXT 1: Gastroenterologist explaining latest colonoscopy results to a patient.

- TEXT 2: Cardiologist describing patients’ clinical history to another physician.

- TEXT 3: Gastroenterologist describing patients’ clinical history to another physician.

- TEXT 4: Pulmonologist describing patients’ clinical history to another physician.

We instructed each participant to record themselves reading these four standardized medical texts aloud using their personal computers and built-in microphones. They were asked to use their natural speaking voice and typical conversational pace to simulate real-world use conditions.

We used two ASR models for transcription, Whisper and WhisperX. (11,12) Whisper is an ASR system trained on extensive multilingual and multitask data from the web, utilizing an encoder-decoder transformer architecture to transcribe speech segments into text. WhisperX enhances Whisper by incorporating voice activity detection (VAD), forced phoneme alignment and diarization methods, providing precise word-level timestamps and speaker attribution. These features make WhisperX particularly suited for detailed analyses required in clinical transcription and video subtitling. Additionally, we implemented a unique approach by chaining WhisperX outputs to GPT-4o for further correction, which we call WhisperX-GPT. This two-step process aims to enhance transcription accuracy without compromising content integrity or overly summarizing the transcripts.

We evaluated the models using two metrics: Levenshtein distance, which measures the number of single-character edits required to match the transcription with the original text, and Word Error Rate (WER), a standard metric that calculates transcription accuracy at the word level as a percentage. To test sensitivity to accented speech, we used one-tailed t-tests to compare the differences between native and non-native groups for each standardized medical text and then for all texts grouped. For the grouped analyses, we normalized all the calculated values using a mean centered approach within each standardized medical text standardized medical text. Finally, we used paired t-tests to compare the performance of non-native groups with (WhisperX-GPT) and without (WhisperX) correction. We conducted all analyses using Python programming language, and the analysis scripts are publicly available on Github (https://github.com/tatonetti-lab/Accent-Project/tree/main).

Additionally, we built an AI engine publicly hosted on the UnityPredict (https://console.unitypredict.com/medical-transcription-engine-with-llm-correction) platform. This engine accepts two types of inputs—audio or transcription files—processes them through WhisperX, then forwards the transcription to GPT-4o for correction, providing users with both the original and corrected transcriptions. This engine is publicly accessible and hosted here.

## Results

We used all 20 audio recordings, consisting of five participants within each subgroup, to assess transcription accuracy. As a baseline, we tested transcription accuracy using the Whisper ASR model across all four participant subgroups for each standardized medical text and for the combined dataset. The model’s performance was evaluated using WER as shown in Figure 1, along with statistical analysis comparing the native and non-native groups presented in Table 1.a. The model performance was significantly better for the native group compared to the non-native group in the combined dataset (WER difference of 11.45, p = 0.003). Supplementary Figure 1, and Supplementary Table 1 present the performance of the model using Levenshtein distance metric.

**Figure 1:**
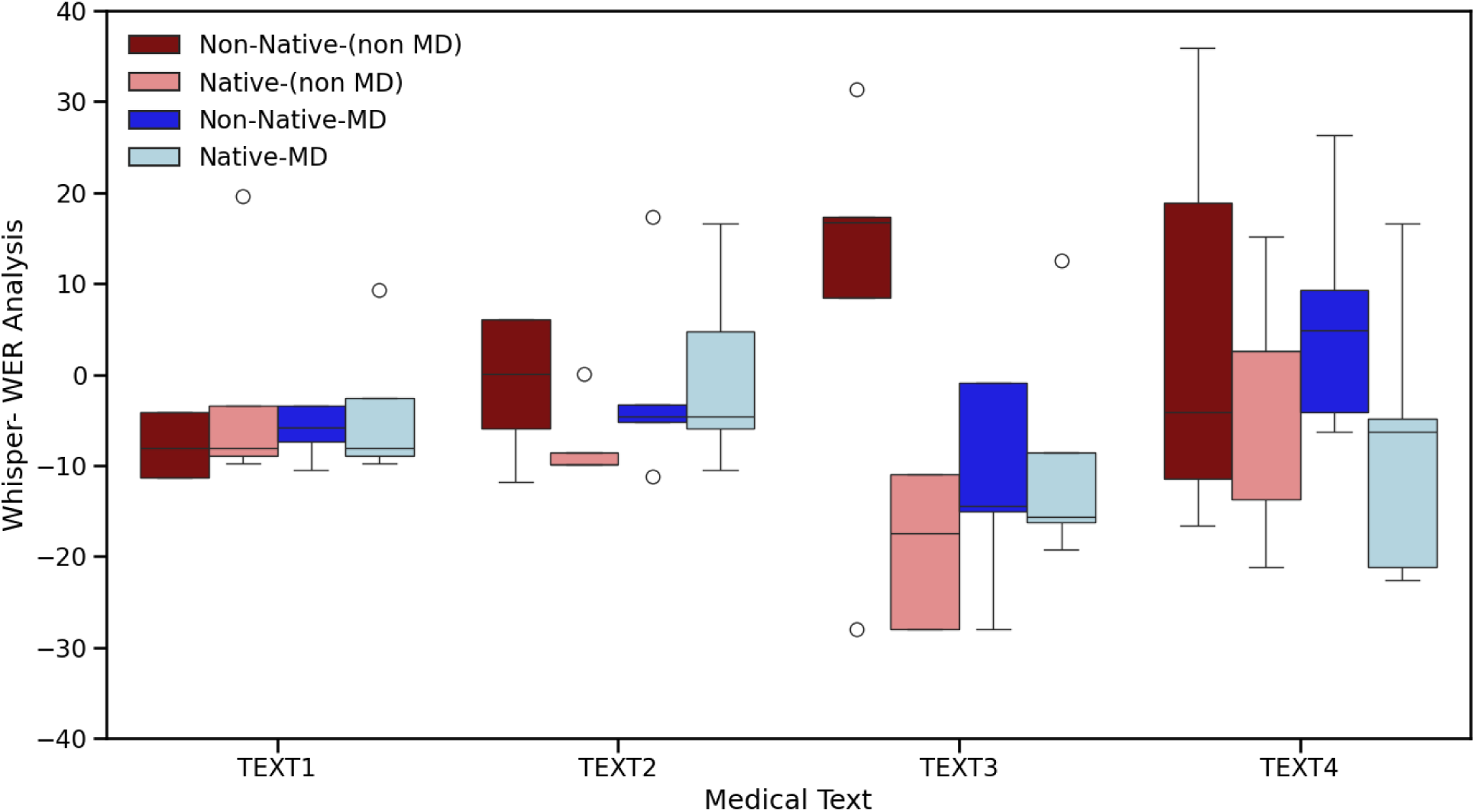
Performance of the Whisper ASR model across four participant subgroups and four Medical Text categories using Word Error Rate (WER) analysis, with values normalized using mean-centered approach within each Text category. Participant groups are defined by accent (Native vs. Non-Native) and medical domain expertise (MD vs. non-MD).

**Table 1:** a) T-test statistical analysis of Whisper performance using WER as the evaluation metric across different text groups and the total dataset, comparing native and non-native groups. The model performed significantly better for the native group than for the non-native group in the total dataset . b) T-test statistical analysis of Whisper-X performance using WER as the evaluation metric across different text groups and the total dataset, comparing native and non-native groups. The model performance was significantly better for the native group compared to the non-native group in the total dataset, indicating higher transcription error rates among non-native speakers.

| a) Whisper (WER analysis) |  |  |  |  |  |  |  |  |
| --- | --- | --- | --- | --- | --- | --- | --- | --- |
| Group - Whisper (WER analysis) | Degree of Freedom (df) | T- statistics | One-tailed- p value | T critical one tail | Accuracy of Group 1 – (Non-Native) | Accuracy of Group 2 –(Native) | Difference | Confidence Interval |
| TEXT1 | 18 | 0.77 | 0.225 | 1.73 | 19.13 | 15.37 | 3.76 | (-20.42, 12.90) |
| TEXT2 | 18 | 1.08 | 0.148 | 1.73 | 22.92 | 15.37 | 7.55 | (-22.26, 7.16) |
| TEXT3 | 14 | 2.65 | 0.009 | 1.76 | 38.82 | 17.21 | 21.62 | (-39.1,-4.14) |
| TEXT4 | 18 | 1.48 | 0.078 | 1.73 | 40.44 | 29.93 | 10.52 | (-25.46,4.42) |
| Total | 60 | 2.85 | 0.003 | 1.67 | 30.33 | 18.88 | 11.45 | (19.43, -3.47) |
| b) Whisper -X (WER analysis) |  |  |  |  |  |  |  |  |
| Group - WhisperX (WER analysis) | Degree of Freedom (df) | T-statistics | One-tailed- p value | T critical one tail | Accuracy of Group 1 – (Non-Native) | Accuracy of Group 2 –(Native) | Difference | Confidence Interval |
| TEXT1 | 18 | 0.80 | 0.217 | 1.73 | 10.4 | 9.1 | 1.35 | (-4.89, 2.19) |
| TEXT2 | 18 | 1.30 | 0.106 | 1.73 | 12.5 | 9.9 | 2.58 | (-6.77, 1.60) |
| TEXT3 | 18 | 2.17 | 0.022 | 1.73 | 19.8 | 14.3 | 5.53 | (-10.89, -0.14) |
| TEXT4 | 18 | 1.78 | 0.046 | 1.73 | 19.5 | 15.3 | 4.15 | (-9.11, 0.76) |
| Total | 78 | 3.22 | 0.001 | 1.66 | 15.5 | 12.1 | 3.40 | (-5.50, -1.30) |

We then applied the same analysis to assess the performance of the WhisperX ASR model. The results using WER are shown in Figure 2, with corresponding statistical comparisons between native and non-native groups presented in Table 1.b. WhisperX demonstrated improved accuracy, reflected by lower error rates across all four medical text groups and the combined dataset, for both native and non-native speakers compared with Whisper. However, a significant difference in performance (WER difference of 3.40, p value = 0.001) between native and non-native groups remained. Supplementary Figure 2 and Supplementary Table 2 present the performance of WhisperX using the Levenshtein distance metric.

**Figure 2:**
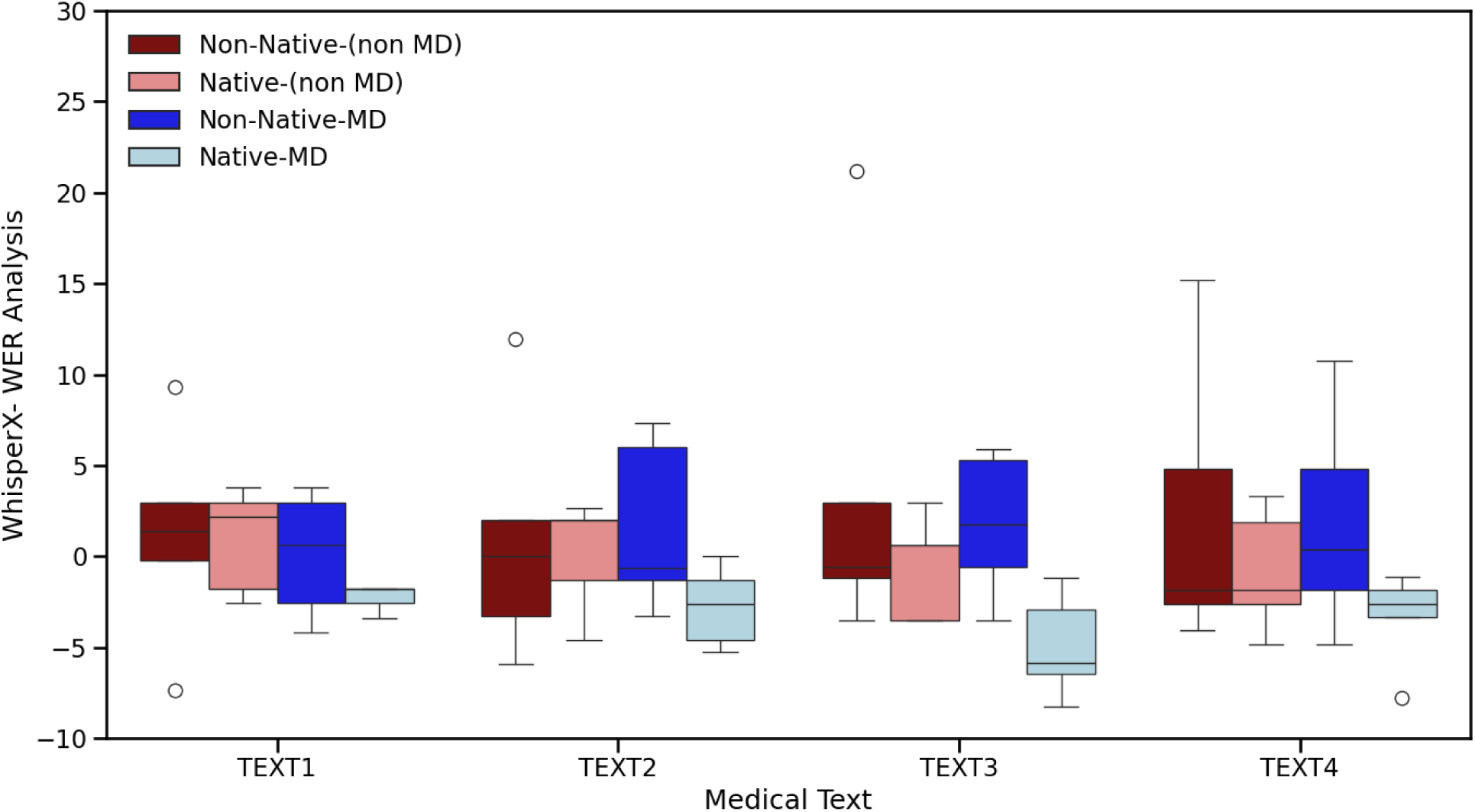
Performance of the Whisper-X,ASR model across four participant subgroups and four Medical Text categories using Word Error Rate (WER) analysis, with values normalized using mean-centered approach within each Text category. Participant groups are defined by accent (Native vs. Non-Native) and medical domain expertise (MD vs. non-MD).

For correction, we passed the transcription results from the WhisperX model to GPT-4o (WhisperX-GPT) and evaluated the performance after correction using both Levenshtein distance and WER metrics. We found that WhisperX-GPT significantly improved the WER for every medical text, with reductions in errors ranging from 4.52 to 6.89 (p<0.001 for all comparisons). Figure 3 illustrates this improvement in performance and Table 2 contains the statistical analysis.

**Figure 3:**
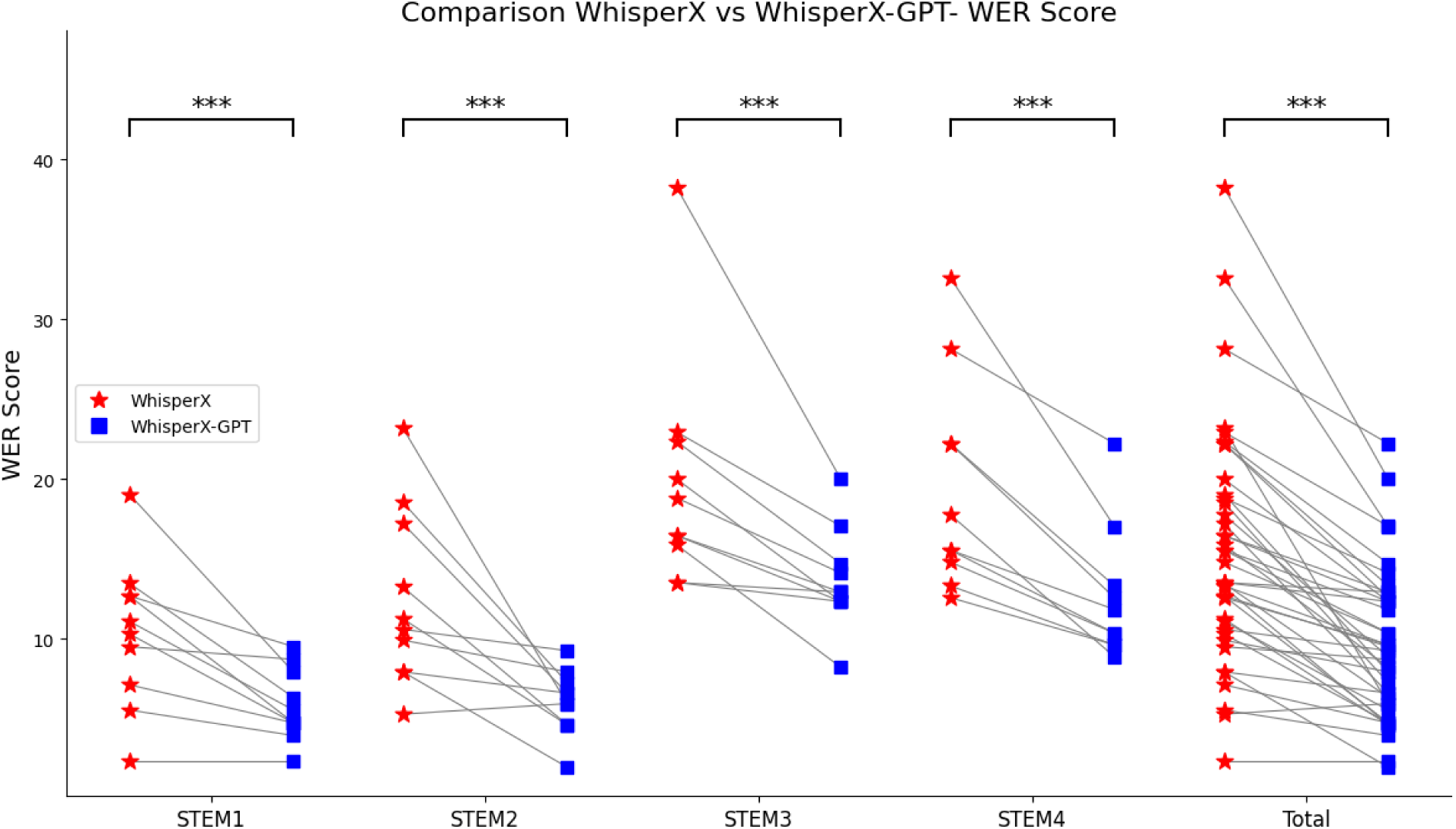
Comparison of Whisper-X and WhisperX-GPT performance using WER scores across non-native speaker groups and four medical text categories. We calculated p-values using paired t-tests, with statistical significance indicated by *** within each group.

**Table 2:** T-test statistical analysis comparing WhisperX and WhisperX-GPT performance using WER as the evaluation metric across non-native groups. The results show significant improvement in transcription accuracy for the WhisperX-GPT model across all medical text groups and in the total dataset compared with the same subgroups of nonnative group.

| Group – Whisper-X vs WhisperX GPT4(WER analysis) | Degree of Freedom (df) | T-statistics | One-tailed-p value | T critical once tail | Accuracy of Group 1(Whisper-X-Non-Native) | Accuracy of Group 2 (Whisper-X-GPT Non-Native) | Difference | Confidence Interval |
| --- | --- | --- | --- | --- | --- | --- | --- | --- |
| TEXT1 | 9 | 4.03 | 0.0015 | 1.833 | 10.397 | 5.87 | 4.52 | (-5.35, 3.69) |
| TEXT2 | 9 | 3.62 | 0.0028 | 1.833 | 12.518 | 6.09 | 6.43 | (-6.82, 6.03) |
| TEXT3 | 9 | 3.90 | 0.0018 | 1.833 | 19.823 | 13.71 | 6.12 | (-7.04, -5.2) |
| TEXT4 | 9 | 5.60 | 0.0002 | 1.833 | 19.481 | 12.59 | 6.89 | (-7.82, -5.96) |
| Total | 39 | 8.43 | 1.27E-10 | 1.685 | 15.555 | 9.57 | 5.99 | (-6.40, -5.58) |

## Discussion

AI systems will become increasingly integrated into healthcare. As they do, automated speech recognition (ASR) models will serve as a critical bridge between users and AI, playing an essential role in ensuring accurate and efficient clinical communication. However, concerns regarding the variable performance of ASR models across diverse user populations, particularly among individuals with different language backgrounds, limit their effectiveness and may contribute to poor clinical outcomes. In this study, we evaluated the transcription accuracy of two ASR models, Whisper and WhisperX, across native and non-native English speakers with and without formal medical training. WhisperX demonstrated superior performance compared to Whisper. However, both models suffered when transcribing accented speech. Implementation of a post-processing pipeline Whisper-X-GPT significantly improved the transcription accuracy, reduced errors, and minimized outliers for non-native English speakers. The correction process narrowed the distribution of transcription performance, suggesting that large language model integration provides a stabilizing effect. This reduction in variability is clinically meaningful, as it minimizes the risk of unpredictable or egregious transcription failures that could compromise clinical documentation or decision-making. In clinical workflows, where precision is paramount and even minor transcription errors can lead to miscommunication, diagnostic inaccuracies, or inappropriate interventions, reducing variability is critical to patient safety. These findings highlight the importance of evaluating not only mean performance metrics but also distributional characteristics when assessing the fairness and reliability of AI tools in healthcare.

Performance metrics such as the Word Error Rate (WER) and Levenshtein distance provide useful benchmarks, however, they may not fully capture clinically consequential errors. In medical contexts, transcription inaccuracies that affect medication names, dosages, procedural terms, or patient instructions may carry disproportionate clinical risk despite contributing minimally to overall error rates. Thus, future evaluations of ASR systems in healthcare should incorporate clinically weighted error assessments that reflect the relative importance of transcription fidelity across different types of content. Moreover, the lack of diverse linguistic representation in model training datasets may exacerbate these issues, underscoring the need for equitable model development and subgroup-specific performance reporting. This study also has other limitations that highlight opportunities for future research. Our sample size was small, limiting statistical power to detect subtle differences across groups. Participants recorded audio using personal devices in uncontrolled environments, introducing variability in sound quality and background noise—factors that, while realistic, may confound performance assessment.

Additionally, our operational definition of native versus non-native English speakers may not fully capture the complexity of linguistic identity or accent variation, and the use of scripted medical texts likely does not reflect the spontaneity and nuance of real clinical communication. As ASR systems become increasingly embedded in real-time clinical documentation, future work should focus on expanding evaluations to larger and more diverse cohorts, incorporating unscripted conversations, and developing domain-specific error classification frameworks. These efforts will be essential to ensure that ASR and LLM technologies are integrated into clinical workflows in a manner that preserves safety, accuracy, and equity.

Nonetheless, our benchmarking approach provides a notable competitive advantage over existing studies by uniquely incorporating accent and medical expertise as evaluation components. To the best of our knowledge, this is the first study systematically examining transcription biases across these dimensions and proposing a practical method of addressing these biases through chaining with additional language models. This methodology is publicly accessible and testable on the UnityPredict platform, facilitating transparency and replicability.

## Conclusion

This study highlights the importance of evaluating and addressing transcription biases in automated speech recognition (ASR) models in clinical settings, particularly those related to speaker accent and domain expertise. We found that WhisperX is a more robust model, particularly when chained with a large language model (GPT-4o) for post-processing, especially for non-native speakers. Moving forward, deploying ASR models in clinical settings will require broader evaluations across more diverse domains, speaker scenarios, and languages. Establishing multi-dimensional evaluation frameworks will be essential to ensure these systems remain reliable and meet the demands of inclusive, high-quality healthcare delivery.

## Data Availability

We conducted all analyses using Python programming language, and the analysis scripts are publicly available on Github.
Additionally, we built an AI engine publicly hosted on the UnityPredict platform. This engine accepts two types of inputs audio or transcription files processes them through WhisperX, then forwards the transcription to GPT-4o for correction, providing users with both the original and corrected transcriptions. This engine is publicly accessible and hosted here.

https://github.com/tatonetti-lab/Accent-Project/tree/main

https://console.unitypredict.com/medical-transcription-engine-with-llm-correction

## Author contributions

Y.F.: Co-conceptualization, co-designed the analyses, performed data collection, performed statistical analysis, co-wrote the manuscript, and approved the final draft.

J.S.S: Co-conceptualization, co-designed the analyses, performed data collection, co-wrote the manuscript, and approved the final draft.

A.K.: Performed data collection, edited the manuscript for important intellectual content, and approved the final draft.

A.S.: Performed data collection, edited the manuscript for important intellectual content, and approved the final draft.

S.F.: Performed data collection, edited the manuscript for important intellectual content, and approved the final draft.

H.L.: Performed data collection, edited the manuscript for important intellectual content, and approved the final draft.

J.B.: Performed data collection, edited the manuscript for important intellectual content, and approved the final draft.

K.T.: Performed data collection, edited the manuscript for important intellectual content, and approved the final draft.

M.Z.: Performed data collection, edited the manuscript for important intellectual content, and approved the final draft.

N.F.: Performed data collection, edited the manuscript for important intellectual content, and approved the final draft.

N.S.: Performed data collection, edited the manuscript for important intellectual content, and approved the final draft.

S.T.: Performed data collection, edited the manuscript for important intellectual content, and approved the final draft.

R.K.: Performed data collection, edited the manuscript for important intellectual content, and approved the final draft.

R.C.: Performed data collection, edited the manuscript for important intellectual content, and approved the final draft.

T.N.: Performed data collection, edited the manuscript for important intellectual content, and approved the final draft.

Y.Y.: Performed data collection, edited the manuscript for important intellectual content, and approved the final draft.

K.H.: Performed data collection, edited the manuscript for important intellectual content, and approved the final draft.

Y.L.: Performed data collection, edited the manuscript for important intellectual content, and approved the final draft.

N.W.: Performed data collection, edited the manuscript for important intellectual content, and approved the final draft.

A.A.: Performed data collection, edited the manuscript for important intellectual content, and approved the final draft.

N.P.T.: Performed data collection, edited the manuscript for important intellectual content, and approved the final draft.

## Supplementary Materials

**Figure 1 Supp:**
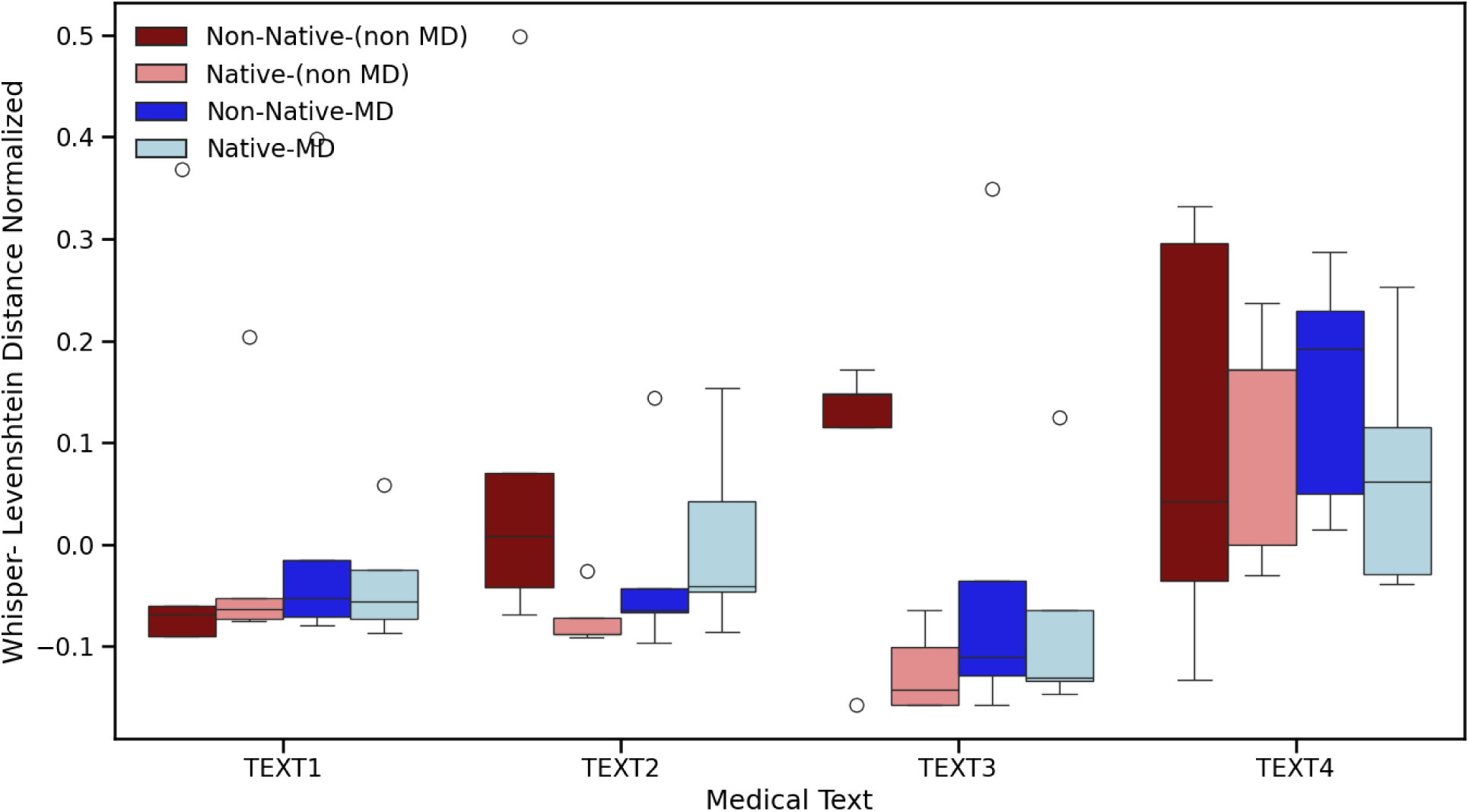
Performance of the Whisper model across four participant subgroups and four Medical Text categories using Levenshtein Distance analysis, with values normalized using mean-centered approach within each Text category.

**Figure 2 Supp:**
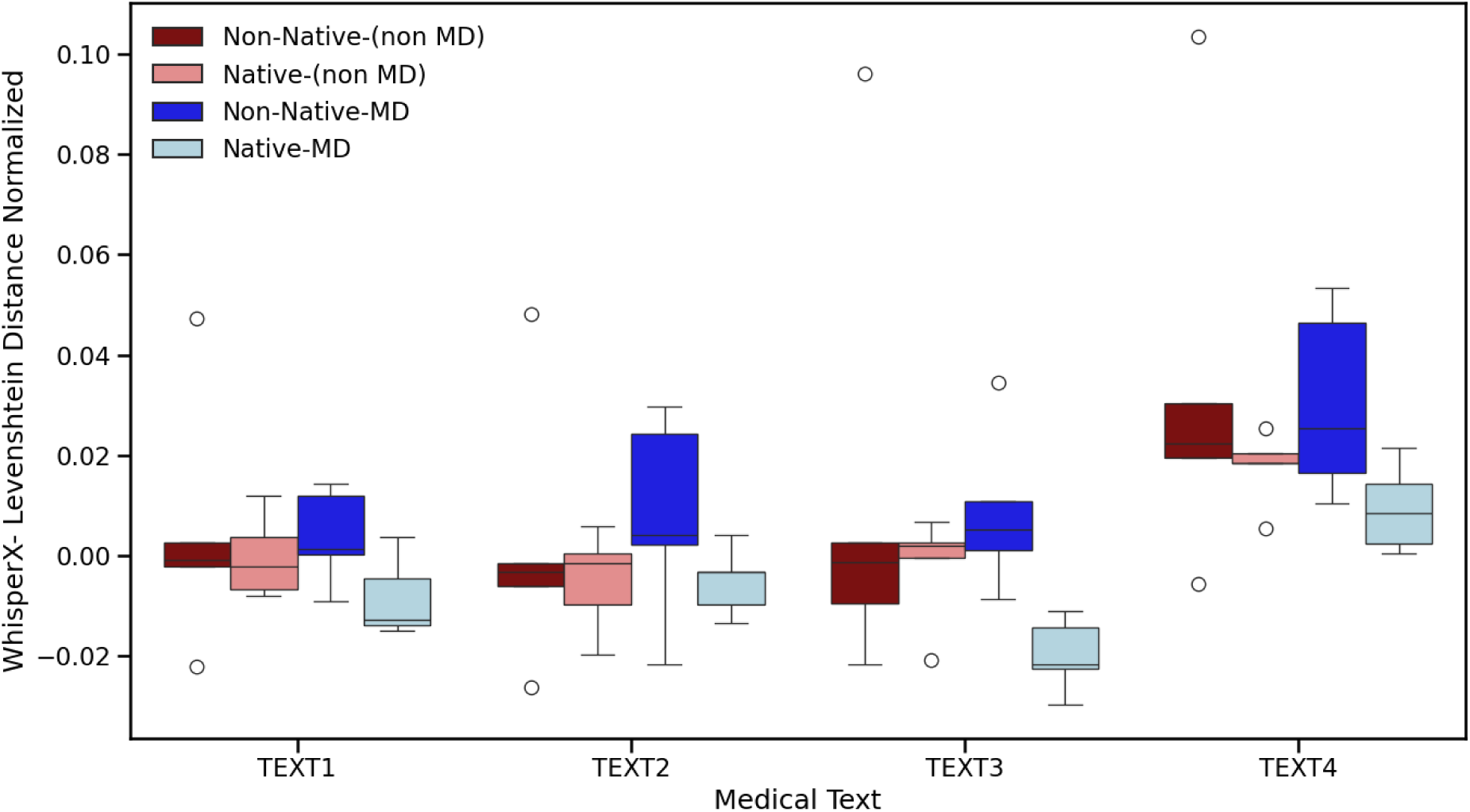
Performance of the Whisper -X model across four participant subgroups and four Medical Text categories using Levenshtein Distance analysis, with values normalized using mean-centered approach within each Text category.

**Figure 3 Supp:**
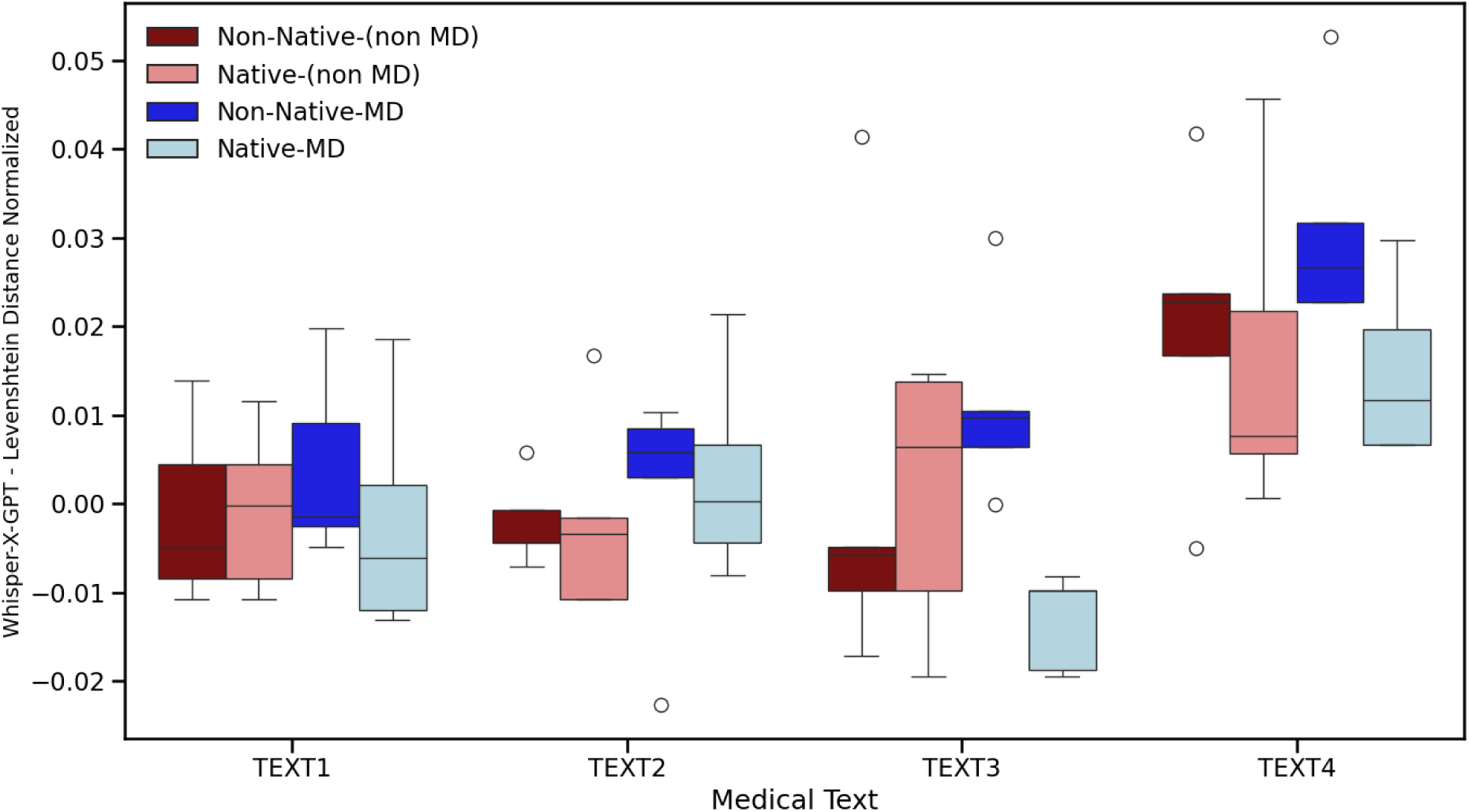
Performance of the Whisper X -GPT4o model across four participant subgroups and four Medical Text categories using Levenshtein Distance analysis, with values normalized using mean-centered approach within each Text category.

**Figure 4 Supp:**
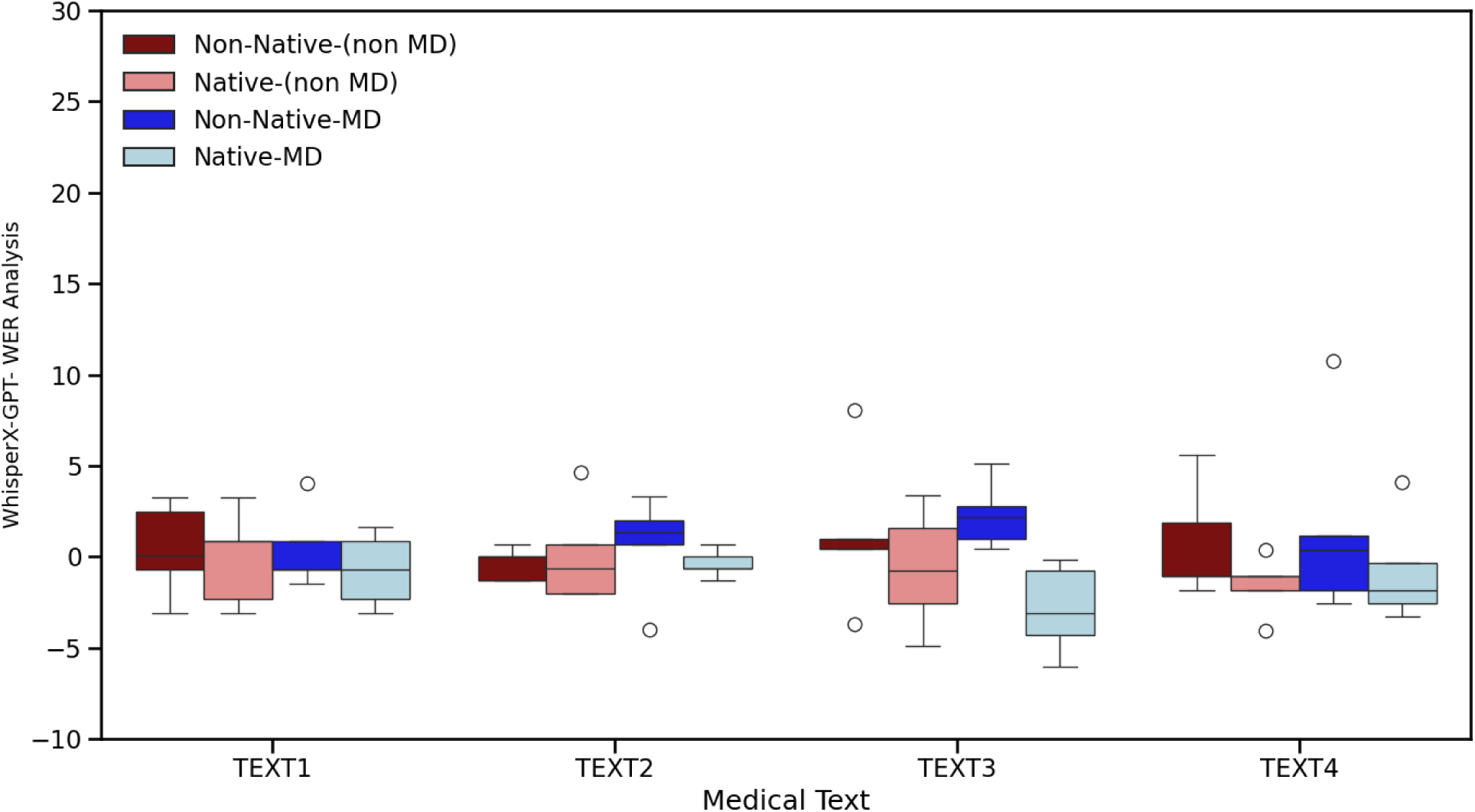
Performance of the Whisper-X+GPT4o, model across four participant subgroups and four Medical Text categories using Word Error Rate (WER) analysis, with values normalized using mean-centered approach within each Text category.

**Table 1 Supp:** T-test statistical analysis of Whisper using Distance as the evaluation metric across different text groups and the total dataset, comparing native and non-native groups.

| Group - Whisper Distance analysis) | Degree of Freedom (df) | T-statistics | One-tailed-p value | T critical once tail | Accuracy of Group 1 – (Non-Native) | Accuracy of Group 2 – (Native) | Difference | Confidence Interval |
| --- | --- | --- | --- | --- | --- | --- | --- | --- |
| TEXT1 | 18 | 0.72 | 0.240 | 1.73 | 0.12 | 0.07 | 0.05 | (-0.19, 0.09) |
| TEXT2 | 18 | 1.10 | 0.142 | 1.73 | 0.16 | 0.09 | 0.07 | (-0.20, 0.06) |
| TEXT3 | 15 | 2.03 | 0.030 | 1.75 | 0.24 | 0.08 | 0.16 | (-0.30, -0.02) |
| TEXT4 | 18 | 0.58 | 0.283 | 1.73 | 0.24 | 0.19 | 0.05 | (-0.18, 0.08) |
| Total | 75 | 2.07 | 0.021 | 1.67 | 0.19 | 0.11 | 0.08 | (-0.14, -0.01) |

**Table 2 Supp:** T-test statistical analysis of Whisper-X using Distance as the evaluation metric across different text groups and the total dataset, comparing native and non-native groups.

| Group - WhisperX Distance analysis) | Degree of Freedom (df) | T-statistics | One-tailed-p value | T critical once tail | Accuracy of Group 1 – (Non-Native) | Accuracy of Group 2 – (Native) | Difference | Confidence Interval |
| --- | --- | --- | --- | --- | --- | --- | --- | --- |
| TEXT1 | 18 | 1.36 | 0.095 | 1.73 | 0.03 | 0.02 | 0.01 | (-0.02, 0.003) |
| TEXT2 | 18 | 1.30 | 0.105 | 1.73 | 0.04 | 0.03 | 0.01 | (-0.03, 0.006) |
| TEXT3 | 18 | 1.93 | 0.035 | 1.73 | 0.05 | 0.03 | 0.02 | (-0.044, 0.004) |
| TEXT4 | 18 | 1.88 | 0.038 | 1.73 | 0.06 | 0.04 | 0.02 | (-0.04, 0.001) |
| Total | 78 | 3.01 | 0.002 | 1.66 | 0.05 | 0.03 | 0.02 | (-0.03, -0.01) |

**Table 3 Supp:**
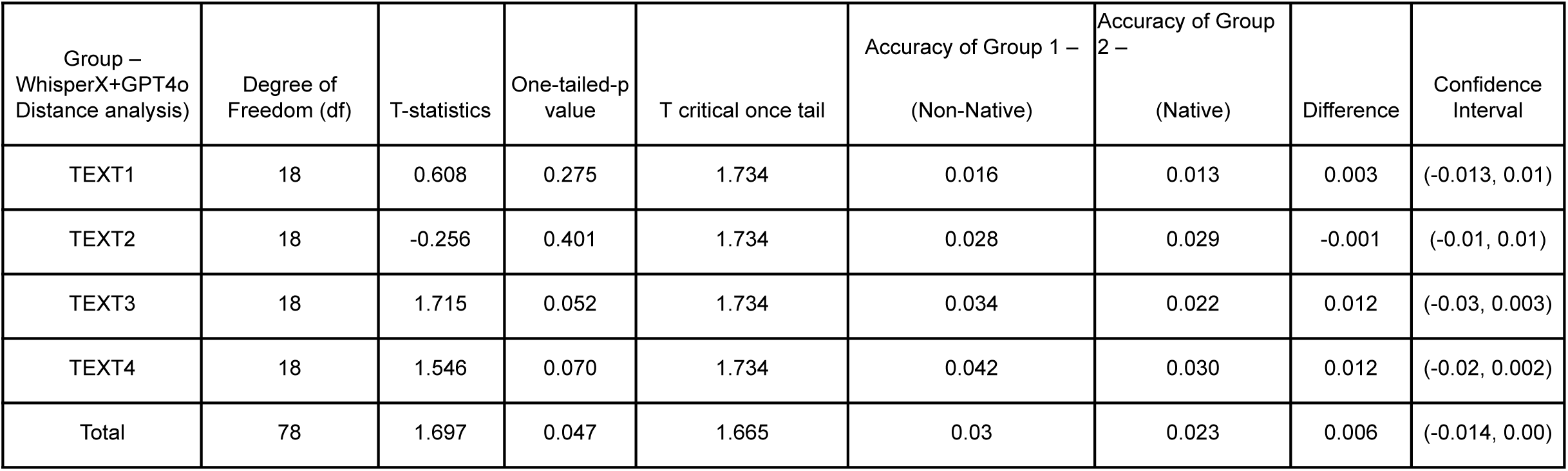
T-test statistical analysis of Whisper-X+ GPT4o using Distance as the evaluation metric across different text groups and the total dataset, comparing native and non-native groups.

| Group – WhisperX+GPT4o Distance analysis) | Degree of Freedom (df) | T-statistics | One-tailed-p value | T critical once tail | Accuracy of Group 1 – (Non-Native) | Accuracy of Group 2 – (Native) | Difference | Confidence Interval |
| --- | --- | --- | --- | --- | --- | --- | --- | --- |
| TEXT1 | 18 | 0.608 | 0.275 | 1.734 | 0.016 | 0.013 | 0.003 | (-0.013, 0.01) |
| TEXT2 | 18 | -0.256 | 0.401 | 1.734 | 0.028 | 0.029 | -0.001 | (-0.01, 0.01) |
| TEXT3 | 18 | 1.715 | 0.052 | 1.734 | 0.034 | 0.022 | 0.012 | (-0.03, 0.003) |
| TEXT4 | 18 | 1.546 | 0.070 | 1.734 | 0.042 | 0.030 | 0.012 | (-0.02, 0.002) |
| Total | 78 | 1.697 | 0.047 | 1.665 | 0.03 | 0.023 | 0.006 | (-0.014, 0.00) |

**Table 4 Supp:**
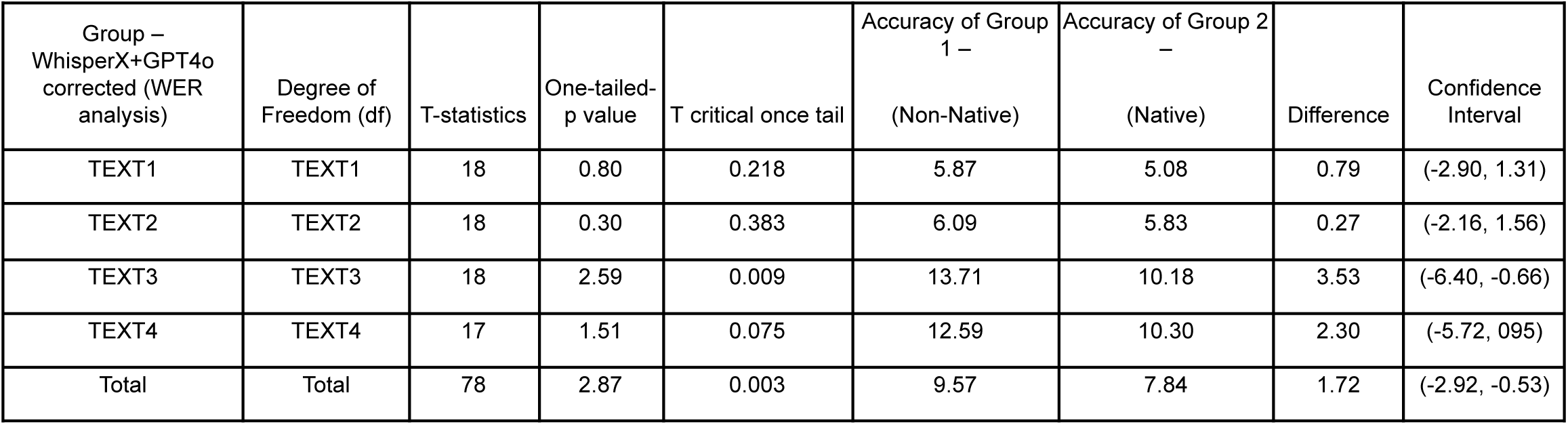
T-test statistical analysis of Whisper-X+GPT4o using WER as the evaluation metric across different text groups and the total dataset, comparing native and non-native groups.

| Group –<br>WhisperX+GPT4o<br>corrected (WER<br>analysis) | Degree of<br>Freedom (df) | T-statistics | One-tailed-p value | T critical once tail | Accuracy of Group 1 –<br>(Non-Native) | Accuracy of Group 2 –<br>(Native) | Difference | Confidence<br>Interval |
| --- | --- | --- | --- | --- | --- | --- | --- | --- |
| TEXT1 | TEXT1 | 18 | 0.80 | 0.218 | 5.87 | 5.08 | 0.79 | (-2.90, 1.31) |
| TEXT2 | TEXT2 | 18 | 0.30 | 0.383 | 6.09 | 5.83 | 0.27 | (-2.16, 1.56) |
| TEXT3 | TEXT3 | 18 | 2.59 | 0.009 | 13.71 | 10.18 | 3.53 | (-6.40, -0.66) |
| TEXT4 | TEXT4 | 17 | 1.51 | 0.075 | 12.59 | 10.30 | 2.30 | (-5.72, 0.95) |
| Total | Total | 78 | 2.87 | 0.003 | 9.57 | 7.84 | 1.72 | (-2.92, -0.53) |

